# Clinical translation of ultrasoft fluorinated elastomer probe for stable, high-density, and bidirectional brain interfaces

**DOI:** 10.1101/2025.04.24.25326126

**Authors:** Jongha Lee, Rongchen Huang, Hyunsu Park, Andrew Spencer, Xian Gong, Matt DeNardo, Foad Vashahi, Florent Pollet, Samantha Norris, Henry Hinton, Méliya El Fakiri, Anant Mehrotra, Julian Bär, Jake Swann, Dave Affonseca, Oliver Armitage, Ryan Garry, Akash Murali, Jordy Tasserie, Caua Fragoso, Romain Albouy, Julia Wang, Elena Grazia Gado, Charles P. Couturier, Angelique C. Paulk, Brian Coughlin, Sydney S. Cash, Beth Costine-Bartell, Benjamin Baskin, Tawny Stinson, Homeira Moradi Chameh, Mandana Movahed, Bamdad Bazrgar, Madeleine Falby, Darren Zhang, Taufik A. Valiante, Ariel Francis, Carlos Candanedo, Ricardo Bermúdez, Jia Liu, Tianyang Ye, Paul Le Floch

## Abstract

Building brain-behavior foundation models requires large functional neural datasets for training, which current implantable Brain-Computer Interfaces (iBCIs) cannot obtain due to the instability of rigid materials in the brain. Achieving high-density neural recordings with wide brain-region access at single-neuron resolution while maintaining long-term stability remains a central challenge. In this study, we present a novel approach to overcome these trade-offs by introducing Fleuron, a fluorinated, ultrasoft, ultra-low-k dielectric materials compatible with thin-film scalable microfabrication techniques enabling scalable, minimally invasive depth electrodes. We successfully integrate up to 1,024 channels within a single Fleuron depth electrode. The combination of novel implant material and geometry enables single-unit level recordings for 18 months in rodent models, and achieves a large number of units detected per electrode across animals. 128-channel Fleuron probes, that cover 8× larger tissue volume than state-of-the-art polyimide counterparts, can track over 100 single-units over months. Stability in neural recordings correlates with reduced glial encapsulation compared to polyimide controls up to 9-month post-implantation. Fleuron probes are integrated with a low-power, mixed-signal ASIC to achieve over 1,000 channels electronic interfaces and can be safely implanted in depth using minimally invasive surgical techniques via a burr hole approach without requiring specialized robotics. Fleuron probes further create a unique contrast in clinical 3T MRI, allowing for post-operative position confirmation. Large-animal and *ex vivo* human tissue studies confirm safety and functionality in larger brains. Neural signals recorded in humans further demonstrate that high-density depth recordings can support decoding of consciousness-related brain states. Together, these results demonstrate the potential of Fleuron probes for clinical translation in iBCIs.

## Main Text

Over 40% of the world’s population suffers from some form of neurological disorder (1), many of which lack effective treatments, severely impacting the quality of life for billions of individuals worldwide. In these cases, implantable Brain-Computer Interfaces (iBCIs) offer significant promise due to their unique ability to decode patient intentions, restore functional capabilities (2, 3), and facilitate personalized therapies by identifying novel biomarkers of neurological diseases. However, to realize widespread adoption, iBCIs must leverage modern artificial intelligence (AI) models to interpret the underpinning dynamics of brain activity involved during behaviors, thoughts, and across patient populations.

However, large AI models typically require immense volumes (*e.g.* billions of data points) of high-quality data for effective training, exceeding by far what wearable technologies and low-resolution iBCIs such as electrocorticography (ECoG) grids and endovascular sensors can currently provide. Recent preclinical studies employing research-grade microelectrode technologies (4) have demonstrated that AI Foundation Models of the brain can be successfully developed using high-density electrophysiological recordings (5, 6). Nonetheless, existing neural interface technologies face significant challenges in clinical translation, primarily due to inherent limitations associated with rigid implant materials. These challenges include (*i*) the trade-off between achieving high bandwidth – often requiring larger probe footprint to support more recording and stimulation channels – while ensuring chronic stability, which is compromised by glial encapsulation (7) and implant micromotion; (*ii*) invasiveness of surgical implantation, which limits clinical usability, and (*iii*) restricted access to deep brain regions critically involved in many neurological disorders (8).

The stability of the brain-electronics interface fundamentally determines the quality of neural data acquired by iBCIs. Downstream processing alone cannot reliably reconstruct latent neural population dynamics from unstable large-scale single- and multi-unit recordings. Yet, capturing these dynamics is essential for decoding brain activity during behavior across diverse patient populations, particularly given intrinsic sources of signal variability such as representational drift (9). State-of-the-art brain-electronics interfaces utilize rigid materials such as silicon and polyimide (10), originally developed for industrial applications rather than optimized for chronic neural integration. This inherent mechanical mismatch imposes significant, yet avoidable, limitations on the scalability and clinical translation of iBCIs.

Here, we address these challenges by introducing a new class of ultrasoft iBCI material: Fleuron. Fleuron polymers mimic more closely the mechanical properties of brain tissues (Young’s modulus ≈ 5 MPa), are compatible with scalable microfabrication, biocompatible and bio-stable, and ultra-low-k (ε = 2.1) thin-film passivation for high-density microelectrode arrays (11, 12). Due to their inherent softness, Fleuron probes maintain stable single-unit resolution despite lateral dimensions eight times larger than those of conventional rigid polyimide probes, effectively minimizing the immune response typically associated with increased probe size. We present various fabricated designs capable of integrating hundreds of sensors and stimulators onto a single implantable thread. Long-term studies demonstrate minimal glial encapsulation around Fleuron probes compared to polyimide controls at 3, 6, and 9 months post-implantation, with stable single-unit recordings maintained for at least 18 months. Chronic recordings using 128-channel Fleuron probes across multiple animals reveal consistently high unit yields and stable performance without significant degradation over 10 months.

We further describe a minimally invasive surgical procedure using an 11-mm inner-diameter burr hole and show that Fleuron probes are visible under clinical 3T MRI at the intended implantation depth without specialized robotic assistance. Acute trials in porcine models confirm the capability of Fleuron probes to record both single- and multi-unit neural activity, while an ovine model demonstrates minimal glial encapsulation after two weeks of implantation. Finally, we demonstrate the feasibility of intraoperative human use of Fleuron probes, enabling high-density single-unit–resolution recordings and robust local field potential responses to a local– global auditory deviant paradigm in anesthetized and awake patients. Leveraging these recordings, we show that neural signals acquired with Fleuron probes support decoding of human consciousness state across subjects, illustrating the potential of stable, high-density depth recordings to capture clinically relevant brain-state dynamics.

### Integration of Fleuron Probes in an iBCI

Clinical applications of iBCIs require integration of multiple technologies such as implantable and hermetic electronics, wireless power and data transmission capabilities, and high-throughput interface to sensors. In this work, we show the integration of Fleuron probes with high-throughput application-specific integrated circuit (ASIC) technology supporting multiplexed amplification and analog-to-digital conversion (ADC) for scaling to multiple thousands of recording and stimulation channels in a proposed implant configuration compatible with minimally invasive burr-hole surgery (**Fig. 1A, B).** Current Fleuron probes can support 128 to 1,024 recording/stimulation channels per thread (**Fig. 1C, D**), where electrode sites’ position and density can be tailored depending on the targeted brain region and implantation depth, due to the compatibility of Fleuron with standard mask- and maskless photo-patterning techniques. Standard metal (Pt, Au, Ti) and metal-oxide (IrO_x_, TiN) frameworks can be used to design high-density interconnects (5-μm pitch) and low-impedance sites. We demonstrate a high-throughput I/O interface connecting Fleuron-encapsulated traces to standard substrates (*e.g.* Si, SiO_2_, Glass), and flip-chip bonded to an ultra-high channel count ASIC (**Fig. 1E, F**), allowing multiplexed neural recording and stimulation from each electrode site.

**Fig. 1.**
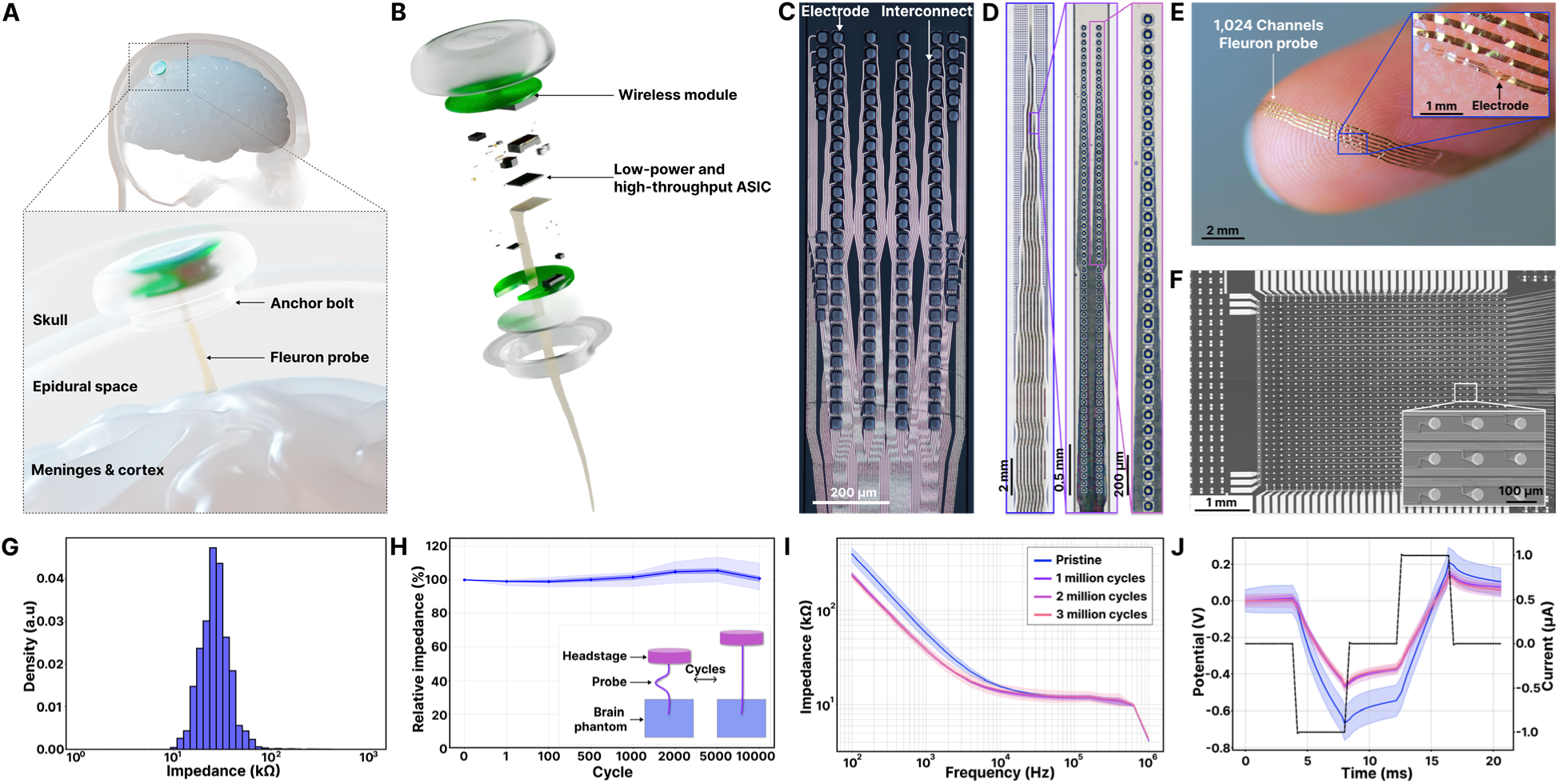
Brain-Computer Interface using Fleuron probes. **(A)** Schematic showing the components of an iBCI using an ultrasoft Fleuron probe. **(B)** Exploded view of (A). **(C)** Stitched bright-field microscopic image of a multi-layer and high-density Fleuron probe with 128 channels and **(D)** 1,024 channels. **(E)** The 1024-channel Fleuron probe placed on a fingertip. Inset shows a magnified view of a region with a high-density of sensors/stimulators. **(F)** Microscope image of the 1024-channel Fleuron probe bonding region to an ASIC with high-density I/O pads. Inset provides a scanning electron microscope (SEM) image of contact pads and interconnect lines **(G)** Distribution of electrode impedance @ 1kHz from 128-channel probes obtained from 10 wafers. **(H)** Normalized impedance distribution for each channel, referenced to the initial value at cycle 0, as a function of the number of bending cycles under a 2 mm bending diameter. Color code: Line = mean; dark blue area = 25^th^ to 75^th^ interquartile range; light blue area = min. to max. values. **(I)** Electrochemical impedance spectroscopy (EIS) characterization and **(J)** Voltage transient (VT) measurement of pristine device, and after 1-, 2- and 3-million stimulation cycles. Color code: Line = mean; dark area = 25^th^ to 75^th^ interquartile range; light area = min. to max. values.

### Electrochemical and Mechanical Performances

Impedance of 32 mm diameter platinum black-coated electrodes across a batch of 10 wafers shows a unimodal distribution with an average impedance value of 39.6 ± 8.0 kΩ at 1kHz across the entire electrode array (**Fig. 1G**), showing the reliability of the multilayer fabrication process using Fleuron in pre-manufacturing stage.

A major limitation of rigid polyimide implants is the progressive displacement, or “drift”, between the probe and surrounding brain tissue over time, driven by the mechanical mismatch and the natural movement of the brain inside the skull. This drift can lead to signal instability, electrical failure, and in severe cases, complete detachment from the implantation site. Fleuron probes inherently mitigate this issue through their extreme mechanical compliance (Young’s modulus ≈ 5 MPa vs. ≈ 3,000 MPa for polyimide), allowing the implant to adapt to biomechanical movements. To assess mechanical stability, we performed cyclic bending tests (**Fig. 1H**), which showed no signs of probe displacement inside the brain phantom, and minimal impedance variations across electrodes – within 10% of the initial impedance value after 10,000 cycles. These findings underscore the suitability of Fleuron probes for chronically stable neural recordings.

Additionally, electrodes embedded in Fleuron can also inject small currents. We show that 32-mm diameter Titanium Nitride-coated electrodes can sustain over three million cycles of stimulation (1 µA cathodic pulse followed by an anodic pulse, each lasting 4 ms, with a 4 ms interpulse delay) with minimal variations in electrochemical impedance and voltage transient (**Fig. 1I, J**), highlighting the technology’s potential for reliable and long-term micro-stimulation applications.

### Minimally Invasive Implantation Process

The headstage and the probe are designed to be implanted and secured using workflows comparable to established minimally invasive neurosurgical procedures, such as implantation of deep brain stimulation (DBS) leads and stereo-electroencephalography (sEEG) electrodes, ensuring its compatibility with existing clinical practice.

**Figures 2A** and **2B** illustrate the implantation workflow in an ex vivo porcine head model. An 11 mm inner diameter burr hole was created, followed by durotomy, after which the Fleuron probe was inserted to the planned depth using a custom 127 μm tungsten stylet. Once the target depth was reached, the stylet was disengaged and carefully withdrawn, the headstage was secured to the skull, and the incision was closed.

**Fig. 2.**
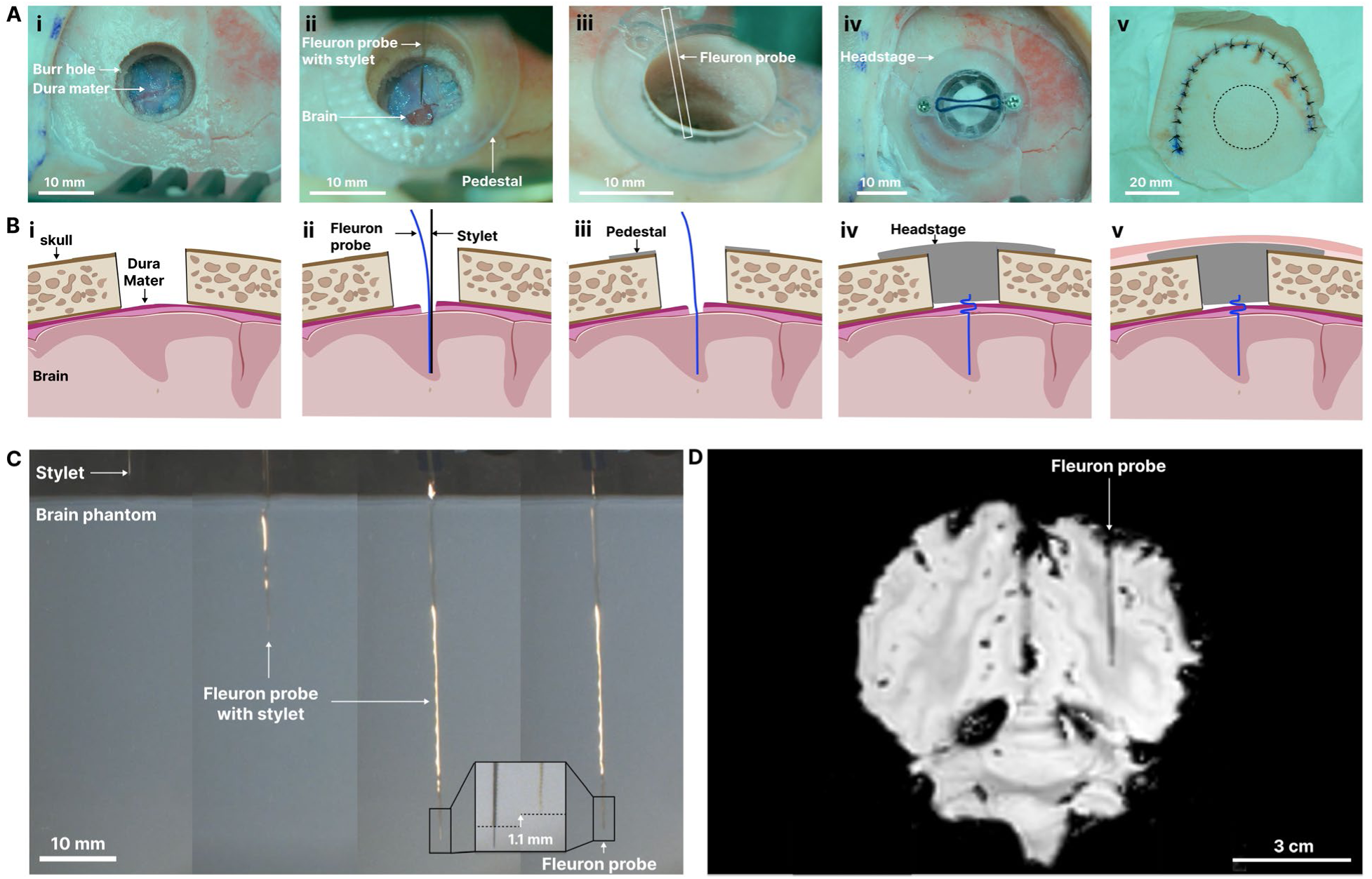
Minimally invasive implantation and imaging of a Fleuron probe for Brain-Computer Interfaces. **(A)** Optical images showing the steps of the minimally invasive implantation procedure of a Fleuron Brain-Computer Interface in an *ex vivo* porcine brain. A standard 14 mm outer diameter and 11 mm inner diameter burr hole is used to access the dura mater. (*i*) Exposed dura mater following burr hole drilling. (*ii*) Implantation of the Fleuron using a stylet after durotomy. (*iii*) Fleuron probe inside brain tissues after the removal of the stylet. (*iv*) Placement of the headstage inside the minimally invasive burr hole. (*v*) Closed surgical site with sutures. **(B)** Corresponding side-view schematic representation of each stage in surgical procedure. **(C)** Optical images of the Fleuron probe with stylet being inserted in a brain phantom (0.6% agarose gel) at 45 mm of depth. Inset shows a magnified view of the probe before and after the retraction of the stylet. **(D)** Magnetic Resonance Imaging (MRI) of the *ex vivo* porcine brain post-implantation.

To quantitatively assess targeting accuracy and deployment stability, implantation was further evaluated in a brain phantom. As shown in **Figure 2C**, the probe was fully inserted to a depth of 45 mm with the stylet in place, demonstrating access to deep subcortical structures. During stylet withdrawal, positional shift was limited to approximately 2%, indicating minimal probe retraction and highlighting the mechanical stability of the Fleuron probe during deployment.

### Magnetic Resonance Imaging (MRI)-Visibility

Depth placement of Fleuron probes can be confirmed after removal of the stylet using MRI. **Figure 2D** shows T2-weighted MRI scans at 3 Tesla from a porcine cadaver model, in which the implanted probe appears as a distinct ^1^H signal void. This MRI visibility arises from Fleuron’s low protium density and its sufficiently large geometrical dimensions, offering a clear contrast against surrounding brain tissue. In contrast, conventional hydrocarbon-based microelectrode arrays, such as polyimide probes, typically lack MRI visibility due to their higher hydrogen content and narrow cross-sectional dimensions.

The visibility of Fleuron probes in post-operative MRI enables confirmation of device placement, which is critical for accurately targeting deep cortical and subcortical structures implicated in many neurological disorders (13).

### Minimized Glial Encapsulation and Tissue Damage Around Larger Probes

State-of-the-art laminar arrays fabricated from conventional rigid materials such as polyimide or silicon often induce significant tissue scarring over time, contributing to signal degradation and instability of neural recordings (7, 14). To minimize gliosis, the dimensions of rigid polyimide probes are typically constrained to widths below 70 µm and thicknesses under 2–5 µm (10).

However, these size limitations restrict the integration of high-density sensing and stimulation sites, reduce tissue coverage, increase capacitive leakage, and elevate the risk of mechanical failure, ultimately limiting the scalability and performance of neural interfaces.

In our previous work, we demonstrated that Fleuron probes with cross-sectional dimensions of up to 250 × 10 µm² induced significantly less gliosis than rigid epoxy-based counterparts (SU-8) following up to 12 weeks of implantation (12). In the present study, we further increase the cross-sectional dimensions of both the Fleuron probes and a polyimide control to 700 × 10 µm² and assess chronic local tissue responses using immunofluorescence imaging in a rat model. We use a 127 µm diameter tungsten stylet, a clinically relevant size that enables deep brain implantation while minimizing the risk of deflection during insertion. Horizontal brain slices (**Fig. 3A**) revealed a markedly lower neuroinflammatory response surrounding the Fleuron probe compared to the polyimide control (**Fig. 3B**), as evidenced by reduced glial fibrillary acidic protein (GFAP)-positive astrocytes – a widely recognized indicator of brain immune response – which is consistent with our previous findings (12). We then quantified the accumulation of GFAP expression as a function of the distance to the surface of the probe (**Fig. 3C**). The results indicate that the Fleuron probes exhibit less glial encapsulation at 3-, 6-, and 9-months post-implantation compared to the polyimide control. Additionally, we observed that the GFAP accumulation at the vicinity of the implant is qualitatively more isotropic around the Fleuron probe, while polyimide controls tend to have a more directional GFAP halo, perpendicular to the largest surface of the probe.

**Fig. 3.**
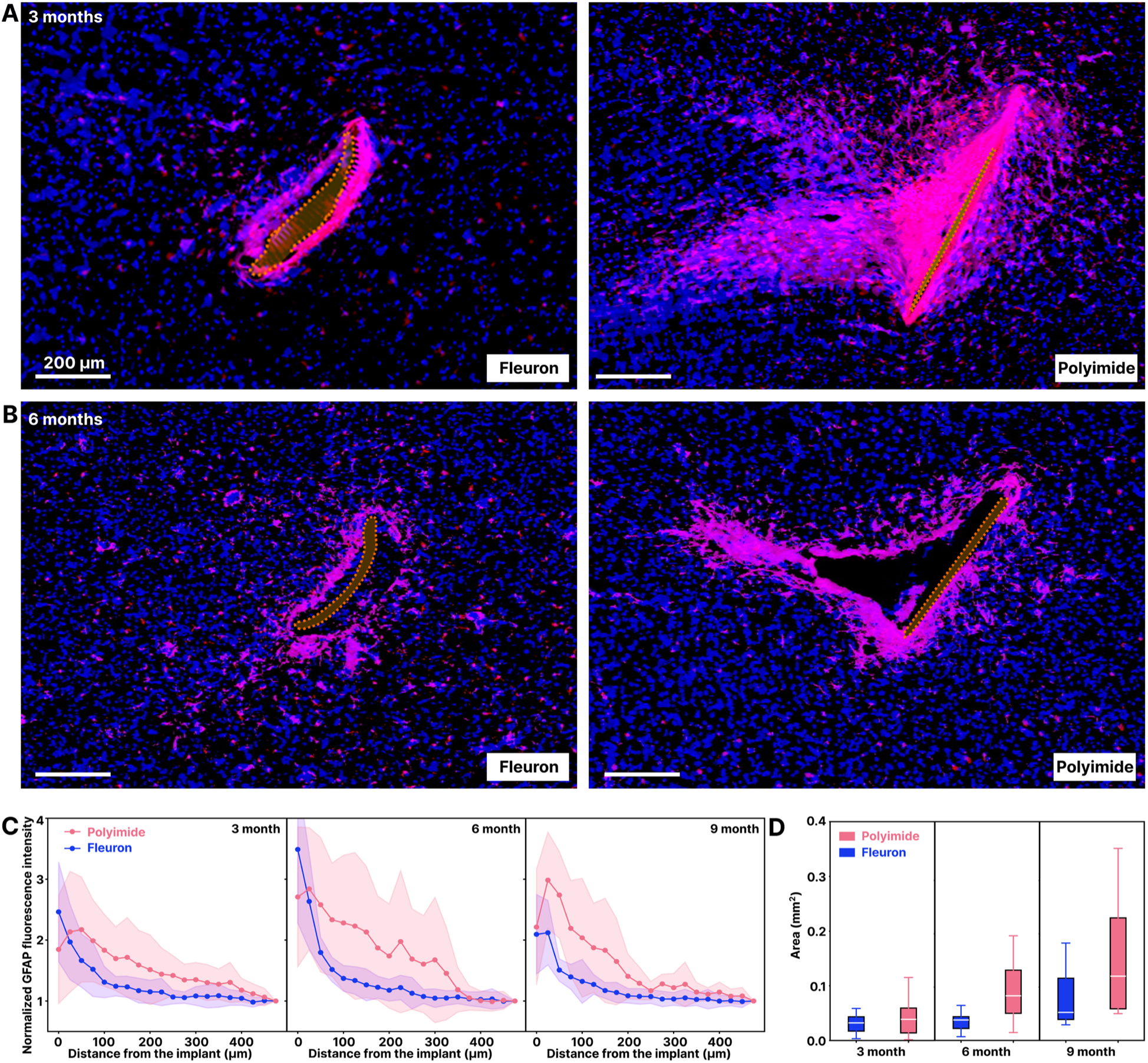
Chronic local tissue response to Fleuron probes versus Polyimide probes. **(A)** Representative immunofluorescence images of horizontal brain tissue sections obtained at 3 months and 6 months following implantation of a Fleuron probe (left) and a polyimide probe (right) in the same rat brain. Staining includes DAPI (blue) for cell nuclei, GFAP (pink) for astrocytes indicating glial scarring, and Iba1 (red) for activated microglia representing neuroinflammatory responses. Probes’ cross-sections are colored in orange. Both probes have identical geometries: 10 µm in thickness and 700 µm in width. Each probe is implanted in one hemisphere of the rodent brain. **(B)** Quantitative analysis of normalized GFAP fluorescence intensity as a function of distance from the probe-tissue interface for Fleuron (blue) and polyimide (pink) implants (N=5, 7 and 8 subjects per time point). Lines show the mean value and shaded regions show the standard error of the mean (SEM). **(C)** Normalized average cavity size in rodents at the electrode-tissue interface for both types of probes.

We then quantify the size of the cavity surrounding the implanted probes (**Fig. 3D**), which reflects both the acute effects of implantation and the cumulative impact of probe micromotion over time. While cavity sizes were similar across groups at 3 months – corresponding to the acute recovery phase – polyimide controls exhibited substantially larger cavities at 6 and 9 months post-implantation, suggesting progressive tissue disruption and instability compared to the Fleuron probes.

Overall, long-term implantation studies show that Fleuron probes, even at larger dimensions, significantly reduce tissue encapsulation and cavity formation compared to rigid polyimide controls. This enhanced biocompatibility enables the integration of a greater number of sensors/stimulators, supporting high-density, tissue-wide, and chronically stable neural interfaces that are difficult to achieve with conventional rigid probes.

### Tissue-wide, High-density Neural Recordings

After establishing a new dimensional baseline for chronically stable brain–electronic interfaces using Fleuron probes (**Fig. 3**), we evaluated the stability of neural recordings in a rat model using 450 µm-wide probes (**Fig. 4A**). Each 128-channel Fleuron probe featured 32 × 32 µm² electrode sites, spaced at 40 µm along the insertion axis, with lateral spacing of either 40 µm or 90 µm between columns, forming six electrode columns in total (**Fig. 4B, C)**. This high-density architecture enables multi-site recordings from individual neuronal units within and across columns to optimize spike sorting performances. The multi-column, high-density structure is specifically designed to capture tissue-wide, non-redundant neural activity while enabling robust spike sorting from individual units across depth and lateral dimensions.

**Fig. 4.**
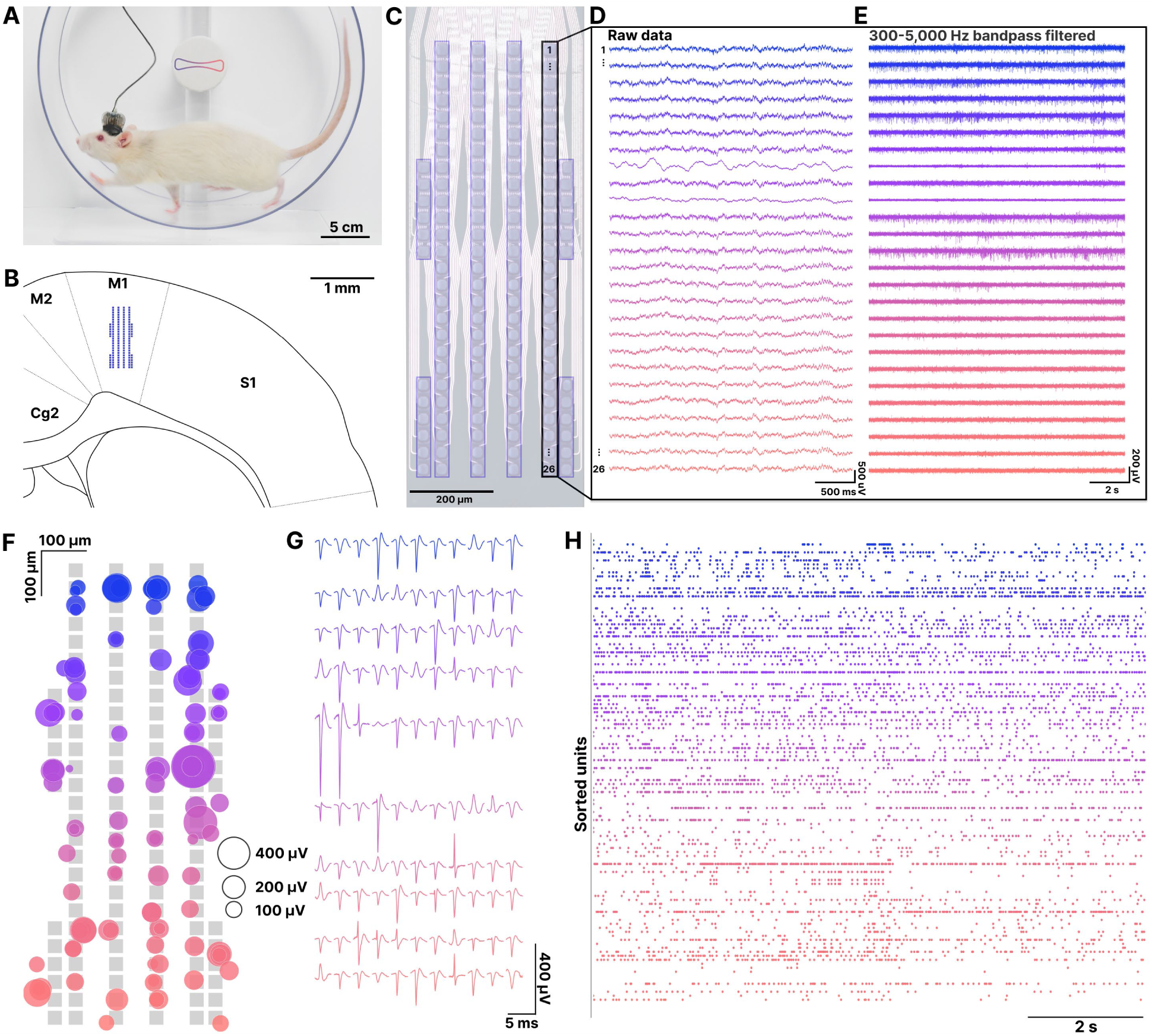
Tissue-wide, high-density single- and multi-unit recordings using Fleuron probes. **(A)** Photograph of a rat implanted with a Fleuron probe. **(B)** Schematic illustration of the probe placement within the primary motor cortex (M1). **(C)** Microscopic image of a 128-channel Fleuron probe with 6 columns of electrodes. **(D)** Representative raw, and **(E)** bandpass-filtered (300–5,000 Hz) voltage traces from a single column of electrodes. **(F)** Visualization of spike sorting results using Kilosort 4, showing the spatial distribution (centroid) and relative amplitude of identified units across the electrodes. **(G)** Average spike waveforms of the sorted units, colored by spatial depth. **(H)** Representative raster plot of all units detected in (G) over a 10-second window.

To evaluate this capability, the probes were implanted in the primary motor cortex (M1, **Fig. 4B**), allowing high-resolution recording of motor-related brain activity during naturalistic behaviors in freely moving rats. Representative voltage traces from a single column highlight the Fleuron probes’ ability to achieve high signal-to-noise ratio (SNR) recordings (**Fig. 4D**, **E**). Rigorous spike sorting using Kilosort 4 (15) revealed that a single Fleuron probe can detect over 100 single units (**Fig. 4F, G**). The spatial centroids and relative amplitudes of the sorted units (**Fig. 4F**), along with their average spike waveforms (**Fig. 4G**) and corresponding raster plots (**Fig. 4H**), collectively demonstrate the ability of Fleuron probes to achieve tissue-wide, high-density, single-unit–level recordings.

### Chronically Stable Single- and Multi-unit level Recordings

To evaluate the long-term performance and recording stability of Fleuron probes, we conducted chronic implantation studies extending up to 18 months in freely behaving rodents. In the longest trial, longitudinal overlays of extracellular spike waveforms reveal consistent spike morphology throughout the 18-month recording period (**Fig. 5A**). To further assess unit stability, high-dimensional spike features were projected into two-dimensional space using Uniform Manifold Approximation and Projection (UMAP), revealing stable clustering patterns over time (**Fig. 5B**). Consistent with these observations, waveform similarity scores across months showed high pairwise similarity of spike shapes throughout the recording period (**Fig. 5C**), suggesting that the same individual neurons or local ensembles were reliably recorded by the same electrodes.

**Fig. 5.**
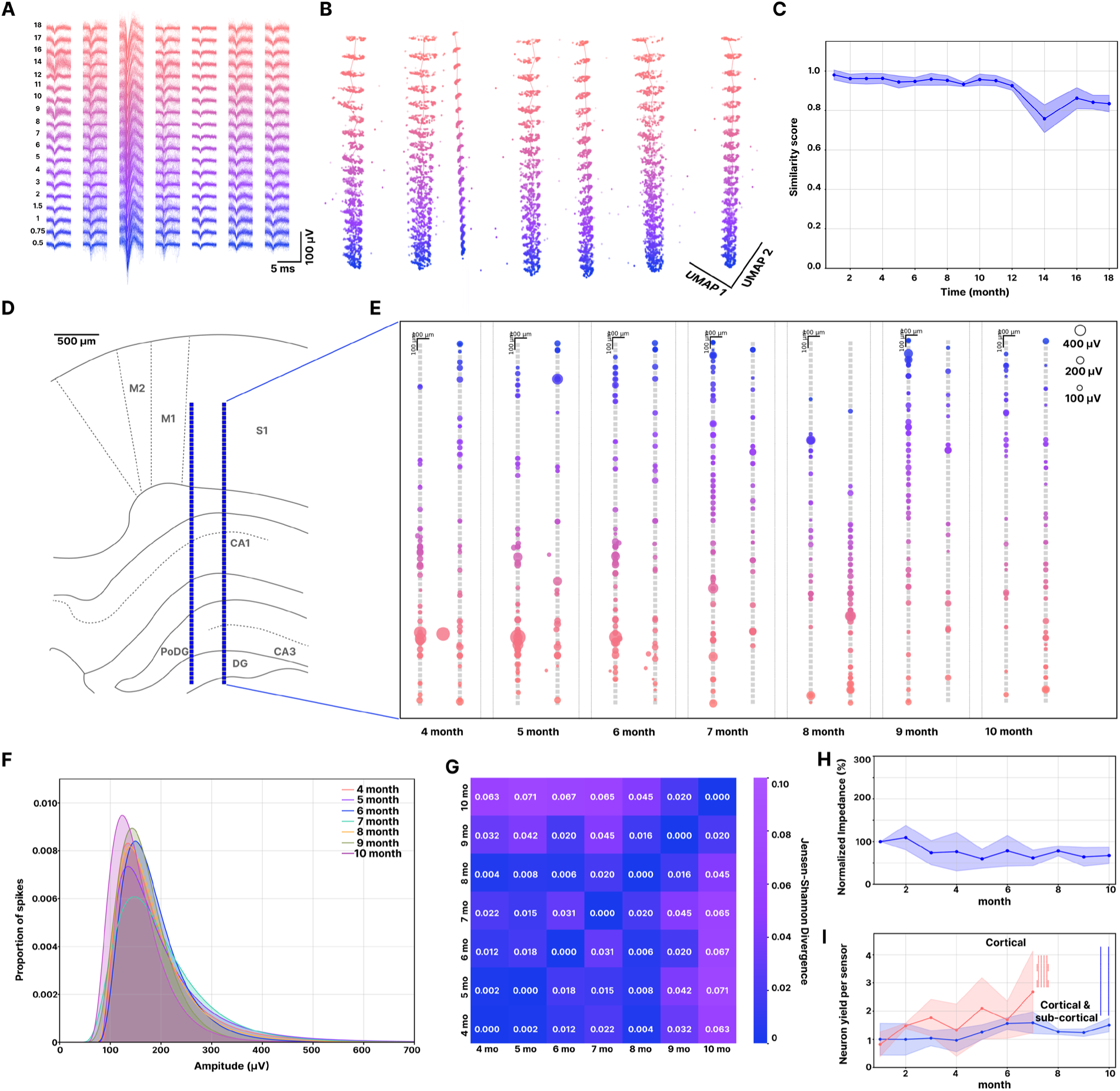
Single-unit level array chronic stability. **(A)** Overlaid detected waveforms recorded from one-month to eighteen-months post-implantation, recorded by a 500 µm wide Fleuron probe. **(B)** Clusters of detected waveforms in two-dimensional Uniform Manifold Approximation and Projection (UMAP) space recorded from one-month to eighteen-month post-implantation. **(C)** Waveform similarity of detected units. **(D)** Rat brain atlas showing the positioning of the probe across cortical and hippocampal layers. **(E)** Amplitude and centroid distribution of detected neurons from 4-, to 10-month post-implantation, sorted by Kilosort 4. **(F)** Distributions of the spike amplitude over time at from 4-, to 10-months post-implantation. **(G)** Jensen-Shannon divergence comparison of each distribution from **(F)** over time. **(H)** Normalized impedance distribution for each channel from 6 implanted devices, relative to the initial value at month 1, as a function of time. Color code: Line = mean; dark blue area = standard deviation. (N = 6 subjects, mean ± standard deviation). **(I)** Average number of neurons detected per electrode (neuron yield) over time (N = 6 subjects, mean ± standard deviation).

To demonstrate stable single-unit recording across varying depths, we designed an extended version of the 128-channel probe featuring two electrode columns and implanted it across multiple cortical and hippocampal layers, including S1, CA1, CA3, and the dentate gyrus (DG) (**Fig. 5D).** In a representative example, we calculated the average waveform amplitude and centroid position of the spike-sorted units using Kilosort from 4 to 10 months post-implantation (**Fig. 5E**). The spatial distribution of active electrodes remained similar over time, and the amplitude distributions of waveform signals were highly consistent across months (**Fig. 5F, G**).

Across the animal cohort (*n* = 6), normalized electrochemical impedance amplitude at 1 kHz (referenced to values measured one-month post-implantation) remains relatively stable over month two to ten (**Fig. 5H**). Notably, we did not observe a drastic drop in impedance over time, which is consistent with bench testing realized on Fleuron probes (12). Additionally, we quantified the average unit yield per electrode across the cohort for both array configurations – six-column arrays targeting cortical regions and two-column arrays for deeper brain structures – and found that unit yield generally increased over month one to ten, further supporting the long-term stability of Fleuron probes in the brain (**Fig. 5I**).

### Translation to Large Animals and *Ex Vivo* Human Tissues

To assess the potential of Fleuron probes for clinical translation, we developed longer threads and implantation shuttles (5–10 cm length) suitable for deployment in large animal models and *ex vivo* human brain tissues. In ovine models, histological analysis performed two weeks after implantation of a 700 µm-wide, 10 µm-thick Fleuron probe revealed no evidence of glial encapsulation at different depth (**Fig. 6A**). To evaluate implantation feasibility in human brain tissue, we performed insertions into *ex vivo* specimens using stained Fleuron probes, comparing acute tissue displacement against commercial stereo-EEG electrodes. Longitudinal brain slices showed that, upon deplantation, Fleuron probes left no residual cavity, in contrast to the stereo-EEG controls, highlighting the minimal invasiveness of the probe and stylet (**Fig. 6B**).

**Fig. 6.**
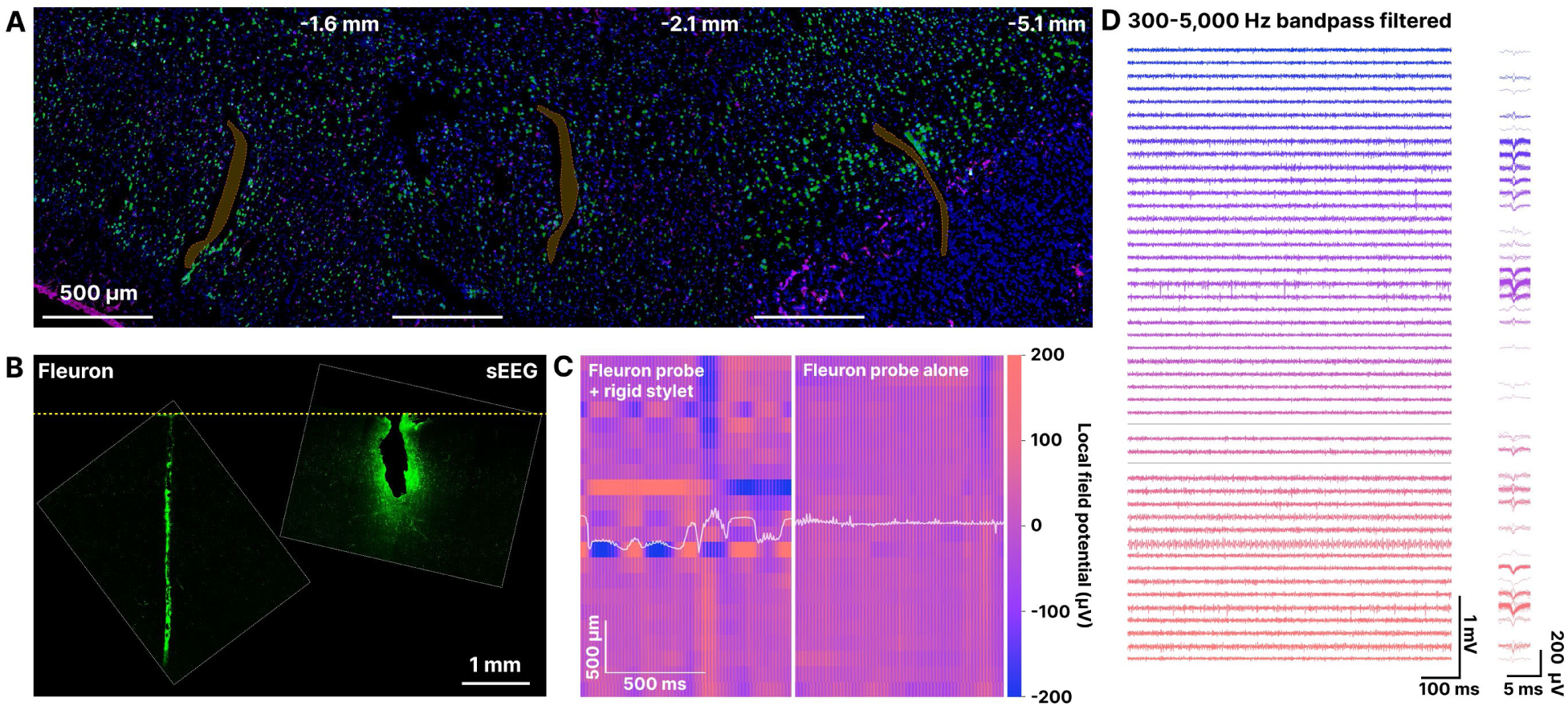
Translation to large animal model and *ex vivo* human tissue. **(A)** Representative immunofluorescence images of horizontal brain tissue sections obtained at 2 weeks following implantation of a 700 µm wide, 10 µm thick Fleuron probes in an ovine model. Staining includes DAPI (blue) for cell nuclei, NeuN (green) for neurons, GFAP (pink) for astrocytes indicating glial scarring, and Iba1 (cyan) for activated microglia representing neuroinflammatory responses. Probes’ cross-sections are colored in orange. **(B)** Representative fluorescence images of vertical brain *ex vivo* human brain slices after implantation and explanation of a stained Fleuron probe (left) and a stereo-EEG probe (right) as control. The dashed yellow line represents the surface of the human tissue sample from which the probes were implanted. **(C)** Local field potential amplitude as a function of time for the Fleuron probe before (left) and after (right) removal of the stylet. Each row represents a different channel, ordered from top to bottom by depth of implantation The white line represent the estimated motion (scale in µm) of the probe relative to the brain (16). **(D)** (Left) Representative filtered voltage traces during acute recordings in porcine model. (Right) Detected single units during the recording.

Lastly, Fleuron probes were evaluated in a porcine model for intraoperative electrophysiological recordings. Using recently developed methods for high-density laminar arrays (16), we assessed the relative motion of the probe before and after removal of the rigid tungsten stylet. The results show that brain pulsation-induced drift is significantly reduced following stylet removal (**Fig. 6C**), highlighting the mechanical stability of the brain-Fleuron probe interface. We also confirmed the ability of Fleuron probes to reliably capture single-unit activity in large animal models (**Fig. 6D**).

### Intra-operative Recordings in Humans

We performed intra-operative neural recordings in human patients undergoing cortical resection surgery to assess the clinical feasibility of the implantation and deplantation procedures developed in animal models.

Implantation trajectory was planned using standard neuronavigation and MRI pre-operative imaging (**Fig. 7A**). Following implantation at approximately 1 cm depth in the cortex, we observe that the Fleuron probe coupled with the natural motion of the brain due to its compliance and the presence of slack between the implantation site and the headstage (**Fig. 7B, C**).

**Fig. 7.**
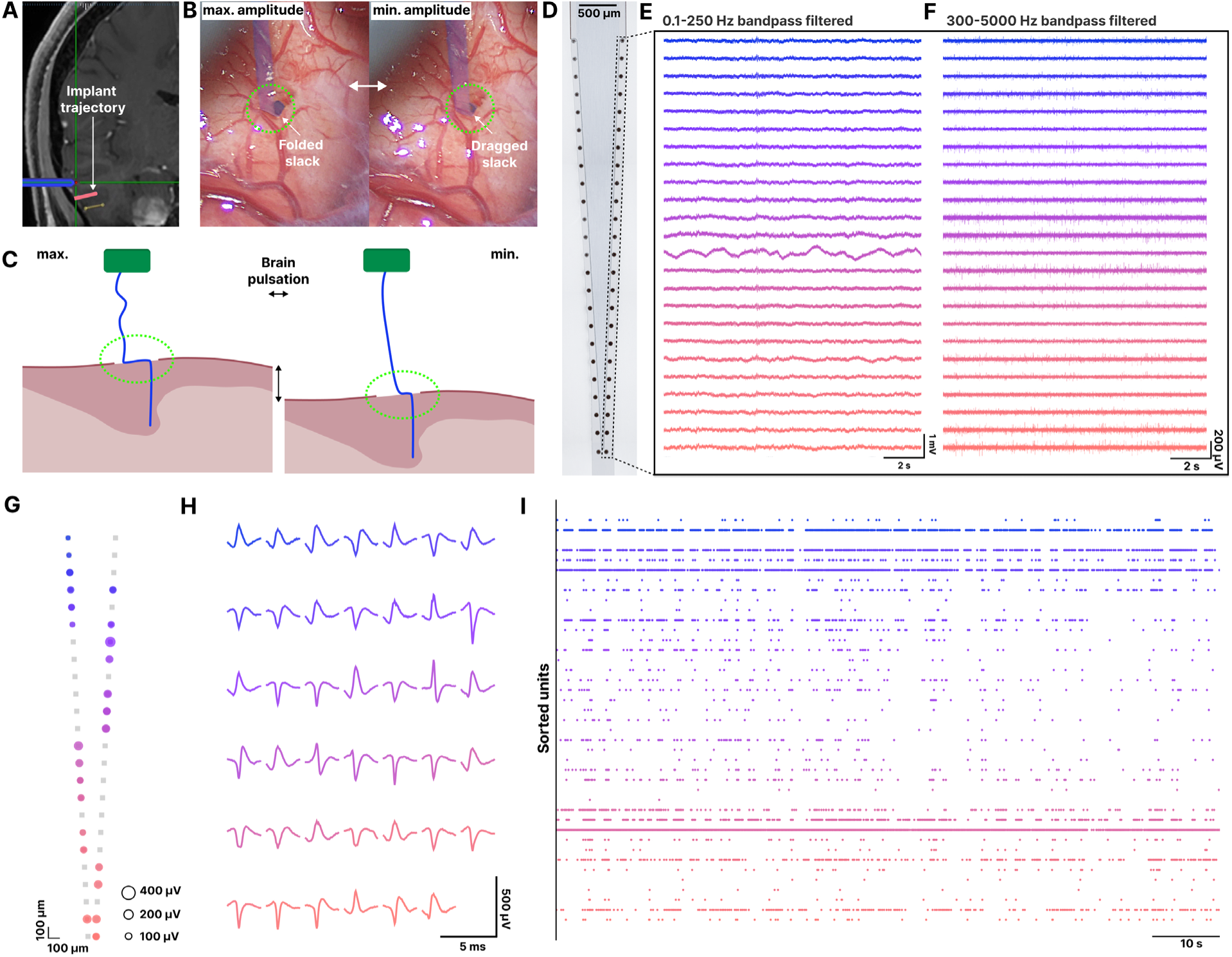
Acute neural recordings using Fleuron probes in human cortex. **(A)** Pre-operative MRI overlaid with the planned implantation trajectory following neuronavigation calibration. **(B)** Intraoperative photographs of a Fleuron probe (colored in blue) on human cortical surface during the cardiac cycle, showing the probe slack displacement between the maximum and minimum of amplitude of brain movement. **(C)** Schematic representation of probe motion induced by the cyclic brain movement. **(D)** Microscopic image of the implanted Fleuron probe, **(E)** Raw and **(F)** 300-5,000 Hz band-pass filtered neural signals acquired from each channel shortly after insertion. **(G)** Visualization of spike sorting results using Kilosort 4, showing the spatial distribution (centroid) and relative amplitude of identified units across the probe’s channels. **(H)** Average spike waveforms of the sorted units, colored by spatial depth. **(I)** Representative raster plot of all units detected in (G) over a 100-second window.

Representative raw and band-pass–filtered voltage traces demonstrate detection of both local field potentials and stable spiking activity from the onset of implantation and throughout the entire intraoperative recording window (∼20 min; **Fig. 7D-F**). Spike sorting revealed the presence of both somatic and non-somatic single units distributed along the length of the probe, indicating stable multi-site single-unit recordings during the procedure (**Fig. 7G–I**).

### State-Dependent Neural Dynamics during Human Intra-operative Recordings

To determine whether intracranial neural signals recorded with Fleuron probes encode human consciousness state, we analyzed intraoperative recordings from five subjects under varying anesthesia conditions (**Fig. 8A**). Two subjects were anesthetized throughout the recording, two were awake, and one transitioned from anesthetized to awake. Four subjects additionally underwent a previously established local–global auditory oddball paradigm (17).

**Fig. 8.**
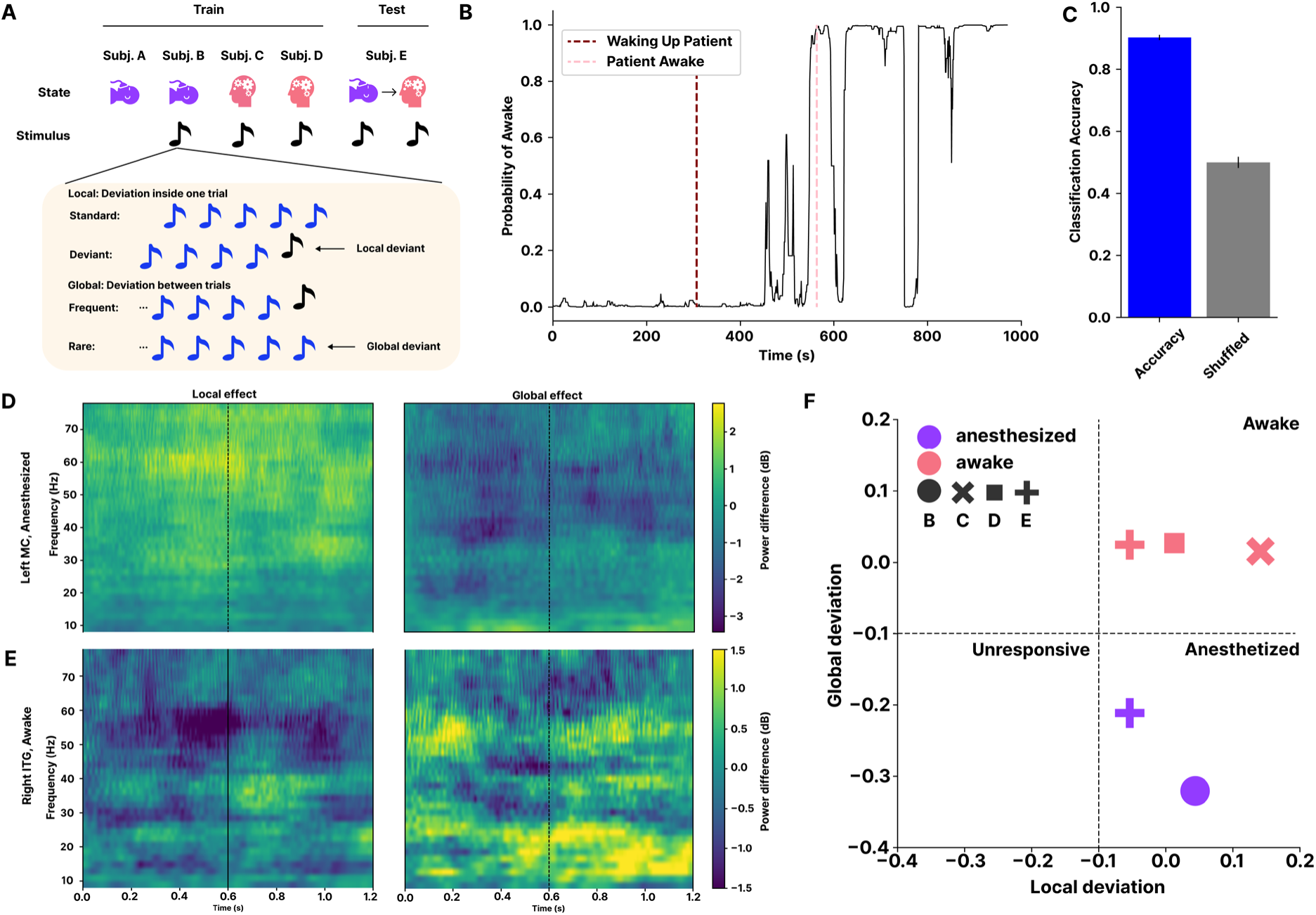
Decoding of human consciousness states from intracranial neural recordings. **(A)** Summary of anesthesia conditions and behavioral paradigms across five human subjects. Two subjects were anesthetized throughout the recording session, two were awake, and one transitioned from anesthetized to awake during recording. Four subjects underwent a local– global auditory oddball paradigm. **(B)** Probability of awake state over time during emergence from anesthesia in a representative subject, decoded using a gradient-boosted classifier trained on local field potential (LFP) features from four subjects and applied to the held-out fifth subject. Behavioral state annotations were determined by anesthesiologist observation. **(C)** Classification accuracy for decoding anesthetized versus awake state compared with shuffled-label controls (n = 30 trials). **(D)** Time-frequency plots of local and global effects recorded from an example channel in the left motor cortex of an anesthetized subject **(E)** and an example channel in the right inferior temporal gyrus (ITG) of an awake subject Time axis is from the start of the first tone, and the dashed line represents the presentation 5^th^ tone. **(F)** Deviation metric computed between standard and deviant-evoked LFP responses for local and global paradigms across subjects and states. The combined local and global deviation metrics differentiate awake, anesthetized, and brain-dead conditions.

A gradient-boosted classifier was trained on LFP features from four subjects and applied to a held-out fifth subject to decode anesthetized versus awake states. In the subject transitioning to wakefulness, the classifier probability of the awake state increased over time and closely tracked behavioral annotations provided by the anesthesiologist (**Fig. 8B**). Across decoding trials, this decoding accuracy reached 89.4% ± 2.3% (n=30) and significantly outperformed shuffled-label controls (49.5% ± 1.7%, n=30), demonstrating robust cross-subject generalization (**Fig. 8C**).State-dependent neural dynamics were further examined using the local–global auditory oddball paradigm, as LFP analyses revealed distinct evoked responses across brain regions and consciousness states. First, time frequency-plots revealed event-related desynchronization and synchronization (ERD/ERS), capturing power changes in neural rhythms that are not strictly phase-locked to stimulus onset. In an anesthetized subject, this ERD/ERS was observed following the local deviant tone, but not the global rare tone (**Fig. 8D**), consistent with residual sensory processing under anesthesia (17). In contrast, recordings of an awake subject exhibited time-ERD/ERS response following both local and global deviants (**Fig. 8E**), consistent with neural signatures associated with conscious processing (17).

To quantify these effects, we computed a deviation metric comparing standard and deviant-evoked responses. The combined local and global deviation metrics robustly differentiated awake, anesthetized, and brain-dead conditions across subjects (**Fig. 8F**). Together, these results demonstrate that intracranial neural recordings obtained intraoperatively with Fleuron probes support decoding of human consciousness state and capture state-dependent cortical processing.

## Conclusion

In this work, we demonstrate that ultrasoft Fleuron probes enable stable, high-density brain– computer interfaces, effectively overcoming the long-standing trade-off between recording density and chronic stability associated with rigid materials like polyimide and silicon. Across rodents, large-animal models, and humans, Fleuron probes support scalable integration with low-power, mixed-signal, bidirectional ASICs and can be deployed at depth using established minimally invasive neurosurgical techniques. Long-term recordings reveal sustained single-unit stability for up to 18 months and robust performance at high channel counts.

Importantly, we demonstrate the first intraoperative use of Fleuron probes in humans, enabling stable, high-resolution single-unit and local field potential recordings at depth within the brain. These recordings capture state-dependent cortical responses to auditory oddball paradigms and support decoding of human consciousness state across subjects. Together, these results establish the clinical translation of Fleuron probes as a platform for long-term, large-scale neural recordings and capable of capturing behaviorally and clinically meaningful brain-state dynamics in humans.

## Acknowledgments

We thank all the clinical study participants, clinical and non-clinical team members, for being involved in this work. Human subject research was conducted at The Panama Clinic with clinical study development done in collaboration with Bioaccess®. The porcine study was supported by NIH NICHD R01 HD099397 to BCB.

## Author contributions

Conceptualization: PLF^1^, TY^1^, JL^1^, JL^1,10^.

All authors from Axoft, Inc. contributed to experimental design, data analysis, and manuscript preparation.

Porcine Study: ACP^3^, BC^3^, SSC^3^, BCB^4^, BB^4^, TS^4^

Ex vivo Human Tissue study: HMC^5^, MM^5^, BB^6^, DZ^7^, TAV^5,7^

Clinical Study Prinicipal Investigator: RB^9^ was the Principal Investigator for the clinical study.

Clinical study surgery: RB^9^, AF^9^ and CC^9^

Manuscript Drafting: JL^1^, RH^1^, PLF^1^

## Ethical Considerations

All experiments involving animals were conducted in accordance with institutional and federal guidelines, and regulations for the care and use of laboratory animals. All rodent procedures were approved by the Institutional Animal Care and Use Committee (IACUC) at Mispro Biotech, Cambridge, MA (Protocol No. 2022-AXO-01). All porcine procedures were approved by the Institutional Animal Care and Use Committee (IACUC) at the Massachusetts General Hospital, Boston, MA (Protocol No. 2010N000198). All ovine procedures were approved by the Institutional Animal Care and Use Committee (IACUC) at Tufts Comparative Medicine-Preclinical Services, Boston, MA (Protocol No. B2023-71). All *ex vivo* human tissue procedures were authorized by the University Health Network Research Ethics Board. The study involving human participants was approved by the Ethics Committee at The Panama Clinic, registered under ClinicalTrials.gov ID <u>NCT06673264</u>. Participants were scheduled to undergo a planned surgery for brain tissue resection of either a tumor or of an epileptogenic lesion. The Axoft device was implanted into normal brain tissue scheduled for surgical resection. All participants provided written informed consent prior to participation.

## Data and materials availability

Sample datasets in the present study are available upon reasonable request to the authors.

## Supplementary Materials

### Materials and Methods

#### Fabrication of Fleuron Encapsulated Neural Probes

Fleuron encapsulated neural probes were fabricated using multilayer thin-film microfabrication processes adapted from fluorinated elastomer-based neural probes reported previously (12). All photoresists and developers were obtained from Kayaku Advanced Materials unless otherwise noted.

Released probes were die-attached to custom printed circuit boards, wire-bonded to PCB pads, and the wire-bonded region was encapsulated with a UV-curable adhesive (NOA 81).

#### Characterization of probe properties

##### Mechanical characterization

Cyclic mechanical testing was performed using a uniaxial tensile testing system (Mark-10) to evaluate the mechanical and electrical stability of Fleuron neural probes under repeated axial loading. Probes were implanted into agarose gel serving as a brain tissue–mimicking medium using a rigid insertion shuttle and electrically connected to an Intan recording system for real-time impedance monitoring. Following implantation and shuttle removal, uniaxial cyclic displacements (±1 mm, 100 cm min⁻¹) were applied along the probe axis for up to 10,000 cycles. Electrode impedance was measured at defined cycle intervals to assess electrical stability as a function of cyclic mechanical fatigue.

##### Electrochemical impedance spectroscopy (EIS)

EIS measurements were performed using a three-electrode setup as described in previous work (12). For each measurement, three frequency sweeps were conducted from 1 MHz down to 100 Hz. A sinusoidal voltage of 100 mV peak-to-peak was applied. Five logarithmically spaced points per frequency decade were measured. For each data point, responses to ten consecutive sinusoids (offset by 10% of the period duration) were accumulated and averaged.

##### Voltage Transient (VT) Measurements

Voltage transient measurements were performed using biphasic current pulses consisting of a cathodic pulse followed by an anodic pulse, each with a duration of 4 ms and separated by a 4 ms interpulse delay. The stimulation amplitude was set to 1 µA. Voltage responses were recorded during stimulation, and electrode polarization voltage was extracted from the transient waveform to assess charge injection behavior and stimulation safety.

#### Animals and Surgery Procedure

Female Sprague Dawley rats (10–12 weeks, Charles River Laboratories) were used in this study. Animals were housed at 20–26 °C, 30–70% humidity, under a 12 h light/dark cycle, with food and water ad libitum.

For implantation, rats were anesthetized with 4–5% isoflurane (induction) and maintained at 1–2%. Stainless-steel screws placed in the frontal bone and cerebellum served as ground electrodes. A 2 × 2 mm² craniotomy was performed in the right hemisphere (M1: AP 0–2 mm, ML 1–3 mm; HIPP: AP –2 to –4 mm, ML 1–3 mm), and the dura was removed. The Fleuron probe, mounted on a releasable shuttle, was implanted using a stereotaxic micromanipulator. At the target depth, the shuttle was retracted. Custom head plate was fixed with C&B Metabond (Parkell) and reinforced with dental acrylic. All procedures followed NIH animal care guidelines and were approved by the Mispro IACUC (protocol AXO-2022-01).

#### MRI of Fleuron probes in Ex Vivo Porcine Heads

Ex vivo porcine heads were implanted with Fleuron probes with micromanipulator. Heads were scanned on a 3T MRI system ([Philips, MR 7700]) using T2-weighted sequences with a slice thickness of 1 mm, optimized for implant visualization. Probe position was identified based on characteristic signal voids along the shaft and tip and visualized in the coronal plane for qualitative assessment of implantation trajectory.

#### Immunohistochemistry

Immunohistochemistry was performed following established protocols with minor modifications. Animals were anesthetized and transcardially perfused with phosphate-buffered saline followed by 4% paraformaldehyde. Brains were extracted with the implanted probes left in situ and post-fixed prior to downstream processing. Tissue sectioning, immunostaining, and imaging were performed by a commercial histology service provider (iHisto).

Brain sections were immunolabeled for neuronal nuclei (NeuN), astrocytes (GFAP), and microglia (Iba1), and counterstained with DAPI. Confocal fluorescence imaging was performed using standardized acquisition parameters across samples.

For quantitative analysis, fluorescence intensity was measured in regions adjacent to the probe tract and normalized to a control region located 500 µm away from the probe, where tissue was assumed to be unaffected by implantation.

#### Electrophysiological Data acquisition

Neural signals were amplified and digitized with a custom system integrated with RHX Data Acquisition Software (Intan Technologies). A custom-designed PCB was used to interface the probe with recording hardware. For rodent experiments, recording was initiated 1 week post-implantation. Neural data was acquired in awake rats, either freely moving or running on a wheel. For porcine and human recordings, data acquisition was performed intraoperatively. Each recording session lasted around 20mins. Recordings were acquired at 20 kHz. Electrode impedance was measured at 1 kHz. Spike detection and sorting were carried out with Kilosort 4.

#### Spike sorting

Data was high-pass filtered at 300Hz, and spike sorting was performed using Kilosort 4 (*18*). The initial automated sorting results were then curated using in one of two ways. At first, curation was done manually in the Phy GUI (https://github.com/kwikteam/phy). Once a number of recordings had been performed and curated thus, a machine learning classifier was trained on existing curated data to perform curation on future spike-sorting results, using the UnitRefine (*19*) implementation in the Python package SpikeInterface. Subsequent curation was performed using this trained classifier to assign quality labels to each cluster based on a range of computed quality metrics.

#### Local Field Potential Data Preprocessing

To obtain the local field potential (LFP) band of the neural signal, raw signal was downsampled to 500 Hz and bandpass filtered to between frequencies of 0.5 Hz and 300 Hz, with notch filters applied at 60 Hz harmonics.

#### Anesthesia State Classification

Frequency information of LFP was obtained per channel using continuous wavelet transformation for 40 log-distributed frequencies between 0.5-50 Hz. The spectrogram was then averaged over channels for each subject, and averaged over sliding 30 second windows. A Gradient Boosted Classifier was then trained on data from subjects A-D to classify anesthetized vs awake, and tested on unseen subject E. Accuracy was compared to a classifier trained on shuffled labels, and reported over 30 stimulations.

#### Local/Global Auditory Oddball Paradigm and Analysis

A local/global auditory oddball paradigm was implemented as described previously (17). Briefly, tones of two frequencies (A and B) are played in 5 tones (trials). Within each trial, the first four tones were identical, and the fifth tone was either the same (e.g., AAAAA) or different (e.g., AAAAB), corresponding to local standard and local deviant trials, respectively. In each block, a sequence of two trial types is played randomly at 80/20 ratio, with the 80% representing a global frequent, and the 20% representing a global rare (i.e. 80% AAAAB and 20% AAAAA). Four block types are played throughout the experiment (frequent AAAAA, rare AAAAB; frequent AAAAB, rare AAAAA; frequent BBBBB, rare BBBBA; frequent BBBBA, rare BBBBB).

Time–frequency representations were computed from LFP data as the difference between multi-taper wavelets of standard and deviant (local effect) or frequent and rare (global effect), baseline corrected with the average power per frequency in the 200 ms before the first tone. As an additional metric to quantify local and global deviation effects, LFP power time courses were first averaged across trials for each condition (local standard, local deviant, global frequent, global rare) within a 0–600 ms window following the onset of the fifth tone. The local deviation metric was defined as the negative Pearson correlation coefficient between the average power time courses of local standard and local deviant conditions. Similarly, the global deviation metric was defined as the negative Pearson correlation coefficient between the average power time courses of global frequent and global rare conditions.

